# Cost-utility of a new psychosocial goal-setting and manualised support intervention for Independence in Dementia (NIDUS-Family) versus goal-setting and routine care: economic evaluation embedded within a randomised controlled trial

**DOI:** 10.1101/2024.08.24.24312530

**Authors:** Abdinasir Isaaq, Claudia Cooper, Victoria Vickerstaff, Julie A. Barber, Kate Walters, Iain A. Lang, Penny Rapaport, Vasiliki Orgeta, Kenneth Rockwood, Laurie T Butler, Kathryn Lord, Gill Livingston, Sube Banerjee, Jill Manthorpe, Helen C. Kales, Jessica Budgett, Rachael Hunter, J Hoe

**Affiliations:** Research Department of Primary Care and Population Health, Wolfson Institute of Population Health, Queen Mary University London, London, UK; Centre for Psychiatry and Mental Health, Wolfson Institute of Population Health, Queen Mary University London, London, UK; Department of Statistical Science, University College London, London, UK; Division of Psychiatry, University College London, London, UK; University of Exeter Medical School, Exeter, UK; Division of Geriatric Medicine, Dalhousie University, Halifax, Nova Scotia, Canada; Faculty of Science and Engineering, Anglia Ruskin University, Chelmsford, UK; Centre for Applied Dementia Studies, University of Bradford, Bradford, UK; Faculty of Medicine and Health Sciences, University of Nottingham, Nottingham, UK; The Policy Institute at King’s, King’s College London, London, UK; Geller Institute of Ageing and Memory, School of Biomedical Sciences, University of West London, London, UK (Prof J Hoe PhD); University California Davis, California, USA

**Author notes:** **Correspondence to:** Prof Rachael Hunter, Priment Clinical Trials Unit (CTU), University College London, 1-19 Torrington Place, London, WC1E 7HB, United Kingdom.

**Keywords:** Dementia, family carer, psychosocial intervention, independence, cost-utility

## Abstract

**Background:** NIDUS-Family is a 6-8 session, psychosocial and behavioural intervention, delivered by non-clinical facilitators, tailored to goals set by dementia-unpaid/family carer dyads. It is effective in terms of attainment of personalised client goals. We aimed to determine if it is cost-effective.

**Methods:** This cost utility and cost-effectiveness analysis is within a two-armed, single masked, multi-site, superiority Randomised Controlled Trial (RCT). We recruited 302 dyads from community settings. Randomisation was blocked and site-stratified, using a 2:1 ratio (intervention: control (goal-setting and routine care)), with allocation by remote web-based system. We calculated the probability that NIDUS-Family is cost-effective for a client with dementia based on Quality Adjusted Life Year (QALY) from health and personal social services and societal perspectives, at £20,000-£30,000 decision thresholds for QALY gained, compared to usual care over 12 months. Analyses were intention-to-treat. **Trial registration** : ISRCTN11425138.

**Findings:** From 30.4.2020-9.5.2022, 204 participants (109 (53.4%) female) were randomised to intervention and 98 (60 (61.2%) female) to control. 218 (72.2%) participants at 6 months and 178 (58.9%) at 12 months provided cost data. There was 89% and 87% probability that NIDUS-Family was cost-effective compared to usual care from personal social services and societal perspectives respectively. Intervention participants accrued on average £8934 (37%) less costs than control participants (95% CI -£59,460 to £41,592).

**Interpretation:** NIDUS-Family is the first personalised care and support intervention to demonstrate cost-effectiveness from the perspective of the quality of life of people with dementia, as well as clinical effectiveness and should be part of routine dementia care.

**Funding:** This work was supported by the Alzheimer’s Society (Centre of Excellence grant 330).

## Background

Around 885,000 people in the United Kingdom (UK) have dementia. Total UK costs for dementia, mostly attributable to social care, will increase from £23.0 billion in 2015, to £80.1 billion in 2040 (1); by 2030, worldwide costs will be an estimated 1.7 trillion US$ (2). Interventions to improve wellbeing and manage symptoms in people with dementia could be highly cost-effective and reduce inequalities in treatment and care, if they reduce long-term care costs by extending the time people with dementia can live, as well as possible, in their own homes, and shift expenditure from emergency care and crisis resolution to prevention. Socioeconomic status predicts receipt of care characterised by later diagnosis, less preventative care (3) and more use of antipsychotics and care home moves (4).

Identifying which interventions are cost-effective is important, due to finite resources in health and care services. The National Institute for Health and Care Excellence (NICE) guidelines build on clinical and cost-effectiveness evidence to inform treatment decisions in the UK. NICE dementia guidelines recommend offering group cognitive stimulation therapy and considering other psychosocial group therapies for people with mild to moderate dementia (3). A recent review of non-pharmacological interventions for dementia found strongest cost-effectiveness evidence for Maintenance Cognitive Stimulation Therapy (MCST) (6). Except for MCST, which is manualised and thus can potentially be delivered by staff without clinical training, implementation of therapies is constrained by staff resources, as they are designed to be delivered by trained clinicians. While dementia care interventions that can be personalised are usually more effective (7), to date personalised interventions have required delivery by clinically trained facilitators (8).

The NIDUS (New Interventions for Independence in Dementia Studies)-Family intervention is a novel approach to care, which enables a fully manualised intervention to be tailored around personalised goals. In NIDUS, family carers and people with dementia (dyads) are supported to set personal goals using Goal Attainment Scaling (GAS) (9) based on their priorities and needs. GAS allows participants to set individualised goals with the facilitator that are defined so an independent evaluator can score their attainment. They then receive a structured support intervention, comprising modules mapped to their goals. Because NIDUS-Family is fully manualised, it can be delivered by trained and supervised, non-clinical, facilitators. This delivery mode was used successfully in the START (STrATegies for carers) intervention, an individual, family carer manual-based intervention that was cost-effective with reference to family carer based QALYs, but did not evaluate cost-effectiveness from the person with dementia’s perspective (10).

Our pragmatic Randomised Controlled Trial (RCT) found home-based goal-setting plus NIDUS-Family was more effective than the control condition (goal-setting completed by the research team and routine care) in supporting dyads’ attainment of personalised goals, over one year (11). The primary aim of this economic evaluation is to calculate the probability that the NIDUS-Family intervention is cost-effective over a 12-month period. We calculated the incremental cost per QALY gained with the NIDUS-Family intervention compared to goal-setting and routine care over 12 months from a health and personal social services cost perspective, and used this to calculate the probability that NIDUS-Family was cost-effective relative to control for a range of cost-effectiveness thresholds for one QALY gained. The secondary objective was to calculate the mean incremental cost per QALY gained over 12 months, and probability of cost-effectiveness from a wider societal perspective (additionally including family carer time).

## Methods

### Ethics committee approval

Camden & King’s Cross Research Ethics Committee (19/LO/1667) approved this RCT on 7.1.2020.

### Study design

This is a cost utility and cost-effectiveness analysis conducted within a two-armed, parallel group, single masked, multi-site, superiority RCT. The published protocol (ISRCTN11425138) is available: https://tinyurl.com/NIDUSfamilyprotocol (12). Two substantial amendments to the protocol (approved 07.04.2020 and 19.09.2022) were made. The first, in response to COVID-19 before study commencement allowed for informed consent, outcome measures and intervention delivery to be conducted remotely via telephone or video call. The second added procedures for a process evaluation (13) and pre-implementation study. Additional 18 and 24 month follow ups (ongoing) were also added.

### Participants

As described previously (11), we recruited participants via professionals working in NHS primary and secondary care, the recruitment database Join Dementia Research, Twitter (now X) and newspaper advertisements. We included dyads consisting of people with dementia and a carer where the care recipient had a documented dementia diagnosis of any type and severity and lived in their own home, and the carer was in at least weekly face-to-face or telephone contact and English-speaking. We excluded dyads if either member was in another research study, the care recipient was in the last six months of life, the carer lacked capacity to consent, or the members of the dyad could not identify at least three eligible GAS goals. Gender was self-reported.

### Randomisation and masking

Allocations were obtained through a remote web-based system: www.sealedenvelope.com provided by the PRIMENT Clinical Trials Unit (CTU). Individual randomisation was blocked and stratified by site using a 2:1 allocation ratio (intervention: control). Randomisation status was concealed from researchers completing outcome measures with carers. We could not mask participants or facilitators.

### Procedures

Trained researchers obtained verbally recorded or written informed consent from all participating carers and care recipients with capacity. Where people with dementia lacked capacity, carers completed a consultee declaration form. Due to COVID-19 pandemic-related restrictions, assessments were conducted via telephone or video call, depending on individual preference; from April 2021, when COVID-19 restrictions were lifted, we also offered in-person assessments. Data were collected at baseline, 6 and 12 months.

### Intervention

NIDUS-Family was delivered by university-employed facilitators, without prior clinical training or qualifications. Initial training comprised ten, one-hour sessions, led by team members including a psychiatrist (CC) and clinical psychologists (including PR), and Alzheimer’s Society volunteers. Facilitators attended group supervision with a clinical psychologist every two weeks. Facilitators delivered 6-8 manualised sessions to dyads over six months, by video-call/telephone (in-person when COVID-19 restrictions permitted). Sessions included carers and people with dementia, or the carer alone; facilitators discussed which arrangement was most appropriate to the dyad’s needs ahead of the session. These manualised sessions were followed by 30-minute catch-up telephone/video calls at 2–3-month intervals (at preference of participants), 6-12 months from baseline to review progress towards goals. Further intervention details are published elsewhere (11).

### Controls

All participants received routine care (care from their GP and in some instances local specialist memory services) and completed goal setting (the primary outcome; procedure detailed below) prior to randomisation.

### Outcomes

The original trial primary clinical outcome for clinical effectiveness was carer-rated Goal Attainment Scaling (GAS)(11,15). GAS allows participants to set individualized, measurable goals with the facilitator that are defined so an independent evaluator can score their attainment. The GAS formula transforms GAS ratings into T-scores so that a score of 50 represents the baseline level, a score of >50 demonstrates improvement and a score of <50 indicates worsening (9). Results for GAS and DEMQOL-Proxy are reported in the main paper (11). We report here the outcomes and cost data collected in this cost-effectiveness analysis. The outcomes analysed here were family carer, proxy-rated client QoL on the Dementia Quality of Life Scale, DEMQOL-Proxy (16); and carer’s own quality of life, rated using the CarerQol (17), which was valued using the CarerQol-7D tariff for the UK (18).

### Cost of NIDUS-Family

We calculated the cost of the NIDUS-Family intervention using data on time spent by facilitators in training and supervision with a clinical psychologist, and contacts that facilitators had with people with dementia and carers to deliver the intervention. Cost per hour of contact for facilitators and supervising clinical psychologist were based on university salary costs for the respective grades and included oncosts and overheads. The total cost of training and supervision was divided by the number of participants in the treatment arm to produce a unit cost per participant for training and supervision. We report the client’s mean cost and standard deviation per dyad in the treatment group.

### Other resource use and costs

Information on services used and support received by the person with dementia were collected using an adapted version of the Client Service Receipt Inventory (CSRI) (19), completed by the carer at baseline (before randomisation), 6, and 12 months. On each occasion, the carer reported service use over the previous 6 months. The CSRI asks questions about services used by the person with dementia, including community-based, emergency, inpatient and outpatient hospital services, any community groups or day centres attended and medication use. It also asked about any carer support services accessed. The CSRI covered adaptations made to the participants’ home during the period of data collection and support received with activities of daily living such as cleaning, whether state-funded, paid out-of-pocket, or unpaid. CSRI questions captured carers’ employment, if any, and any time off work taken to care for their relative with dementia or receive support services. Frequency and intensity of service contacts were reported by group and multiplied by unit costs to estimate total health and social care costs. Unit costs used the most recent nationally published sources (Supplementary Materials Table 1). Medication costs were calculated using the British National Formulary (BNF). For clients who died, a 6-month cost of death of £10,455 was included after their last follow-up (20).

The wider societal perspective analysis included out-of-pocket costs, unpaid help from family/friends, carer employment leave to care for their relative, and voluntary care services. The average time spent per month on unpaid help was calculated from responses to the CSRI on unpaid carer time for different activities. Unpaid help was costed using the replacement method, which values unpaid carer time as if it was paid for, based on the hourly cost of a homecare worker. Unpaid carer time off work was costed using the human capital approach using the UK Office for National Statistics’ median cost per day for full or part-time workers for the carer’s category of employment (21).

### Analysis (Full statistical analysis plan Supplementary material)

Analyses were intention to treat. The primary analysis used a non-parametric 2-stage bootstrap to account for the relationship between costs and outcomes but also for facilitator clustering (22).

Quality-Adjusted Life Years (QALYs) were calculated based on family carer responses to DEMQOL-Proxy using the DEMQOL-U-Proxy classification system (23). QALYs were calculated as the area under the curve (24). Clients who died were entered as 0 after their last follow-up time point. Mean utility values and mean unadjusted QALYs from baseline to 12 months were reported for both groups. The mean incremental difference in QALYs and 95% confidence intervals (CIs) were calculated using linear regression adjusting for baseline utilities, site and accounting for therapist clustering in the calculation of the standard errors. This was reported with bootstrapped 95% CI. No discounting was used as the time horizon is 12 months.

Incremental Cost-Effectiveness Ratio (ICER): We reported the mean incremental cost per QALY gained between the NIDUS-Family arm and control arm at 12 months. The cost in the ICER was calculated from self-reported health and social care resources in NIDUS-Family intervention compared to TAU. The cost of the NIDUS-Family intervention was included for the intervention arm only. The mean difference in costs and 95% CIs were based on bootstrapped results from a linear regression model that included baseline costs and site as covariates and accounted for facilitator clustering.

Cost-effectiveness acceptability curve (CEAC) and cost-effectiveness plane (CEP): The bootstrapped means and 95% CIs for costs and QALYs were used to calculate the probability that the NIDUS-Family intervention is cost-effective compared with control for a range of cost-effectiveness thresholds for one QALY gained. A cost-effectiveness plane shows the bootstrapped results. Missing data was handled assuming that data were missing at random, meaning that missingness is based on factors for which we have complete information and are unrelated to the intervention. The observed factor that predicted a missing ICER was education. We imputed DEMQOL-proxy utility score, total health care costs, and wider societal components for each time point for the recommended 50 datasets using chained equations (multiple imputation using chained equations (MICE)) and predictive mean matching with estimates combined using Rubin’s rules(25). Our primary analyses our imputed; we conducted complete case analyses as secondary analyses. Descriptive statistics are based on complete cases only.

We reported the ICER, CEP and CEAC for the NIDUS-Family intervention compared to control at 12 months from a wider societal perspective using the methods described in the primary analysis but including wider societal costs.

Our a priori sample size calculation indicated 297 (198 intervention; 99 control) participants were required to detect a moderate effect size of 0.5 for the primary outcome comparison (GAS) between intervention and control groups at 5% significance level (2-tailed) with 90% power. The calculation included inflation for intervention arm facilitator clustering (intra-cluster correlation coefficient (ICC) 0.05, average cluster size 20) and 15% loss to follow up (23).

## Results

302 dyads were recruited between 30.4.2020 and 9.5.2022. Twenty-one sites recruited on average 14 participants (Standard Deviation (SD) 9.01; Range 3 -31). Figure 1 (Supplementary material) shows the CONSORT diagram. 247/302 (82%) randomised dyads completed the primary outcome. Tables 1 and 2 describe participants’ baseline characteristics.

**Table 1:**
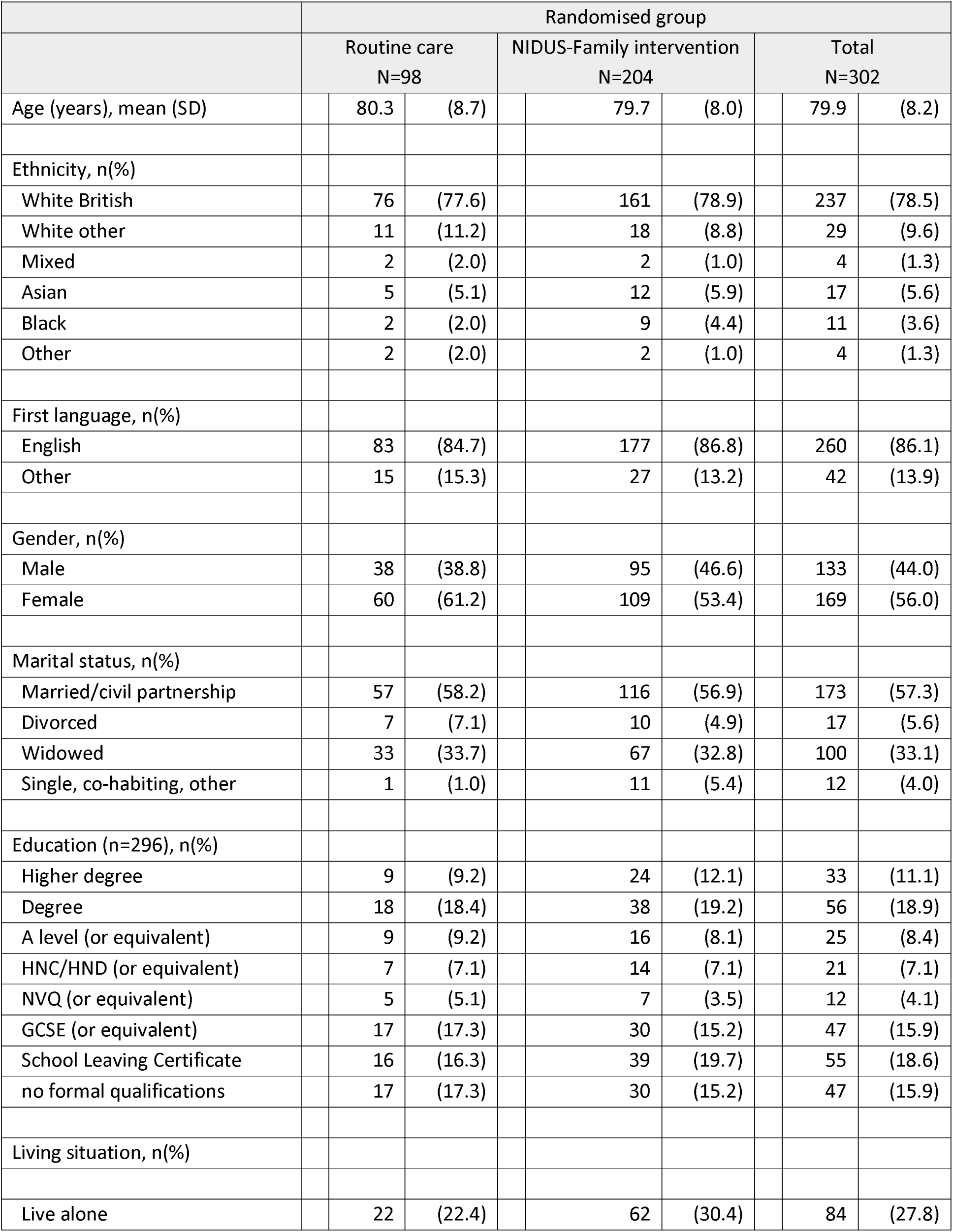
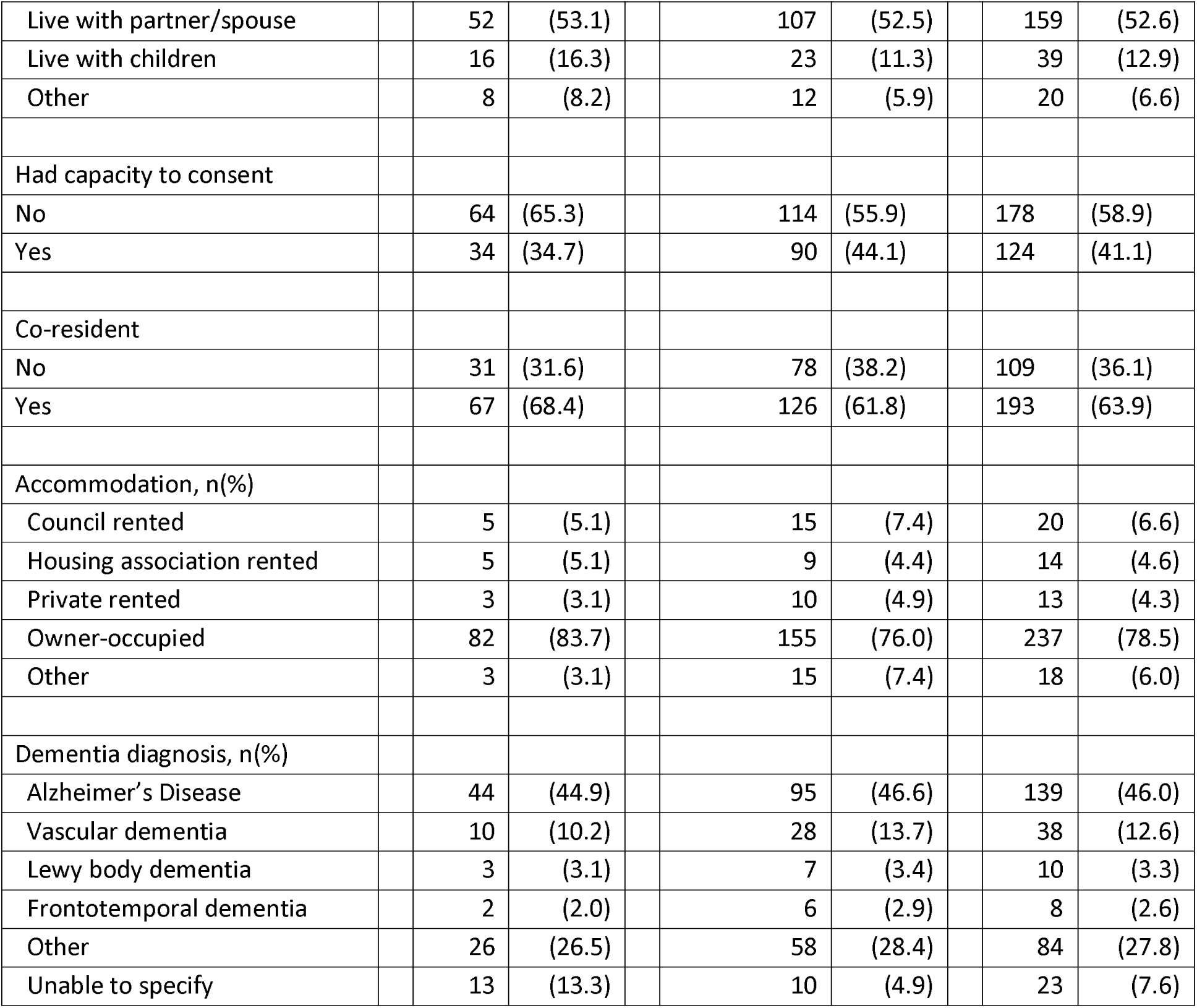
Baseline participant characteristics by randomised group.

| Table 1: Baseline participant characteristics by randomised group |  |  |  |  |  |  |  |  |
| --- | --- | --- | --- | --- | --- | --- | --- | --- |
|  | Randomised group |  |  |  |  |  |  |  |
|  | Routine care<br>N=98 |  |  | NIDUS-Family intervention<br>N=204 |  |  | Total<br>N=302 |  |
| Age (years), mean (SD) |  | 80.3 | (8.7) |  | 79.7 | (8.0) | 79.9 | (8.2) |
| Ethnicity, n(%) |  |  |  |  |  |  |  |  |
| White British |  | 76 | (77.6) |  | 161 | (78.9) | 237 | (78.5) |
| White other |  | 11 | (11.2) |  | 18 | (8.8) | 29 | (9.6) |
| Mixed |  | 2 | (2.0) |  | 2 | (1.0) | 4 | (1.3) |
| Asian |  | 5 | (5.1) |  | 12 | (5.9) | 17 | (5.6) |
| Black |  | 2 | (2.0) |  | 9 | (4.4) | 11 | (3.6) |
| Other |  | 2 | (2.0) |  | 2 | (1.0) | 4 | (1.3) |
| First language, n(%) |  |  |  |  |  |  |  |  |
| English |  | 83 | (84.7) |  | 177 | (86.8) | 260 | (86.1) |
| Other |  | 15 | (15.3) |  | 27 | (13.2) | 42 | (13.9) |
| Gender, n(%) |  |  |  |  |  |  |  |  |
| Male |  | 38 | (38.8) |  | 95 | (46.6) | 133 | (44.0) |
| Female |  | 60 | (61.2) |  | 109 | (53.4) | 169 | (56.0) |
| Marital status, n(%) |  |  |  |  |  |  |  |  |
| Married/civil partnership |  | 57 | (58.2) |  | 116 | (56.9) | 173 | (57.3) |
| Divorced |  | 7 | (7.1) |  | 10 | (4.9) | 17 | (5.6) |
| Widowed |  | 33 | (33.7) |  | 67 | (32.8) | 100 | (33.1) |
| Single, co-habiting, other |  | 1 | (1.0) |  | 11 | (5.4) | 12 | (4.0) |
| Education (n=296), n(%) |  |  |  |  |  |  |  |  |
| Higher degree |  | 9 | (9.2) |  | 24 | (12.1) | 33 | (11.1) |
| Degree |  | 18 | (18.4) |  | 38 | (19.2) | 56 | (18.9) |
| A level (or equivalent) |  | 9 | (9.2) |  | 16 | (8.1) | 25 | (8.4) |
| HNC/HND (or equivalent) |  | 7 | (7.1) |  | 14 | (7.1) | 21 | (7.1) |
| NVQ (or equivalent) |  | 5 | (5.1) |  | 7 | (3.5) | 12 | (4.1) |
| GCSE (or equivalent) |  | 17 | (17.3) |  | 30 | (15.2) | 47 | (15.9) |
| School Leaving Certificate |  | 16 | (16.3) |  | 39 | (19.7) | 55 | (18.6) |
| no formal qualifications |  | 17 | (17.3) |  | 30 | (15.2) | 47 | (15.9) |
| Living situation, n(%) |  |  |  |  |  |  |  |  |
| Live alone |  | 22 | (22.4) |  | 62 | (30.4) | 84 | (27.8) |

|  |  |  |  |  |  |  |  |  |  |
| --- | --- | --- | --- | --- | --- | --- | --- | --- | --- |
| Live with partner/spouse |  | 52 | (53.1) |  | 107 | (52.5) |  | 159 | (52.6) |
| Live with children |  | 16 | (16.3) |  | 23 | (11.3) |  | 39 | (12.9) |
| Other |  | 8 | (8.2) |  | 12 | (5.9) |  | 20 | (6.6) |
| Had capacity to consent |  |  |  |  |  |  |  |  |  |
| No |  | 64 | (65.3) |  | 114 | (55.9) |  | 178 | (58.9) |
| Yes |  | 34 | (34.7) |  | 90 | (44.1) |  | 124 | (41.1) |
| Co-resident |  |  |  |  |  |  |  |  |  |
| No |  | 31 | (31.6) |  | 78 | (38.2) |  | 109 | (36.1) |
| Yes |  | 67 | (68.4) |  | 126 | (61.8) |  | 193 | (63.9) |
| Accommodation, n(%) |  |  |  |  |  |  |  |  |  |
| Council rented |  | 5 | (5.1) |  | 15 | (7.4) |  | 20 | (6.6) |
| Housing association rented |  | 5 | (5.1) |  | 9 | (4.4) |  | 14 | (4.6) |
| Private rented |  | 3 | (3.1) |  | 10 | (4.9) |  | 13 | (4.3) |
| Owner-occupied |  | 82 | (83.7) |  | 155 | (76.0) |  | 237 | (78.5) |
| Other |  | 3 | (3.1) |  | 15 | (7.4) |  | 18 | (6.0) |
| Dementia diagnosis, n(%) |  |  |  |  |  |  |  |  |  |
| Alzheimer's Disease |  | 44 | (44.9) |  | 95 | (46.6) |  | 139 | (46.0) |
| Vascular dementia |  | 10 | (10.2) |  | 28 | (13.7) |  | 38 | (12.6) |
| Lewy body dementia |  | 3 | (3.1) |  | 7 | (3.4) |  | 10 | (3.3) |
| Frontotemporal dementia |  | 2 | (2.0) |  | 6 | (2.9) |  | 8 | (2.6) |
| Other |  | 26 | (26.5) |  | 58 | (28.4) |  | 84 | (27.8) |
| Unable to specify |  | 13 | (13.3) |  | 10 | (4.9) |  | 23 | (7.6) |

### Intervention costs

Dyads received an average of 7 (SD 2) main intervention sessions and 2 (SD 2) catch-up calls. Including 2 hours per dyad of intervention preparation, at £25.51 per hour for facilitator time, the mean cost per dyad of intervention sessions was £238 (SD 69). A clinical psychologist worked two hours a week (for 40 weeks a year for 2.5 years of intervention delivery) to provide training and supervision, at an hourly rate of £58.50 (total cost £11,700), and 10 facilitators each spent an average of 20 hours in training and 20 hours in supervision (£10,204). The mean total cost per dyad for training and supervision was £107.37. Manual printing costs were £5 per participant. Overall, NIDUS-Family intervention mean cost per dyad was £346 (SD 69).

### Resource use

Descriptive statistics for resource use are reported in Table 2 (supplementary material). Costs at baseline, six months and 12 months are reported by allocation group in Table 3. Data to calculate total health and personal social services costs were available for all (n=302) participants at baseline, 207 (68.5%) participants at 6 months and 159 (52.7%) participants at 12 months. Cost of death was imputed for 19 participants for a total of 162 (53.6%) participants with data available to calculate costs across the 12-months.

**Table 2:** Baseline carer characteristics by randomised group.

| Table 2: Baseline carer characteristics by randomised group |  |  |  |  |  |  |
| --- | --- | --- | --- | --- | --- | --- |
|  | Randomised group |  |  |  |  |  |
|  | Routine care<br>N=98 |  | NIDUS-Family<br>intervention<br>N=204 |  | Total<br>N=302 |  |
| Carer Age (years), mean (SD) | 64.0 | (11.5) | 63.1 | (12.9) | 63.4 | (12.5) |
| Carer Ethnicity, n(%) |  |  |  |  |  |  |
| White British | 75 | (76.5) | 157 | (77.0) | 232 | (76.8) |
| White other | 11 | (11.2) | 23 | (11.3) | 34 | (11.3) |
| Mixed | 2 | (2.0) | 2 | (1.0) | 4 | (1.3) |
| Asian | 5 | (5.1) | 11 | (5.4) | 16 | (5.3) |
| Black | 2 | (2.0) | 9 | (4.4) | 11 | (3.6) |
| Other | 3 | (3.1) | 2 | (1.0) | 5 | (1.7) |
| Carer First language, n(%) |  |  |  |  |  |  |
| English | 88 | (89.8) | 189 | (92.6) | 277 | (91.7) |
| Other | 10 | (10.2) | 15 | (7.4) | 25 | (8.3) |
| Carer Gender, n(%) |  |  |  |  |  |  |
| Male | 38 | (38.8) | 52 | (25.5) | 90 | (29.8) |
| Female | 60 | (61.2) | 152 | (74.5) | 212 | (70.2) |
| Carer Marital status, n(%) |  |  |  |  |  |  |
| Married/civil partnership | 77 | (78.6) | 156 | (76.5) | 233 | (77.2) |
| Divorced | 5 | (5.1) | 6 | (2.9) | 11 | (3.6) |
| Single | 9 | (9.2) | 24 | (11.8) | 33 | (10.9) |
| Co-habiting | 4 | (4.1) | 14 | (6.9) | 18 | (6.0) |
| Widowed | 3 | (3.1) | 2 | (1.0) | 5 | (1.7) |
| Other | 0 | (0.0) | 2 | (1.0) | 2 | (0.7) |
| Carer Education, n(%) |  |  |  |  |  |  |
| Higher degree | 18 | (18.4) | 37 | (18.1) | 55 | (18.2) |
| Degree | 30 | (30.6) | 67 | (32.8) | 97 | (32.1) |
| A level (or equivalent) | 11 | (11.2) | 31 | (15.2) | 42 | (13.9) |
| HNC/HND (or equivalent) | 10 | (10.2) | 8 | (3.9) | 18 | (6.0) |
| NVQ (or equivalent) | 4 | (4.1) | 16 | (7.8) | 20 | (6.6) |
| GSCE (or equivalent) | 16 | (16.3) | 26 | (12.7) | 42 | (13.9) |
| School Leaving Certificate | 6 | (6.1) | 7 | (3.4) | 13 | (4.3) |
| No formal qualifications | 3 | (3.1) | 12 | (5.9) | 15 | (5.0) |
| Relationship of carer to person with |  |  |  |  |  |  |
| dementia, n(%) |  |  |  |  |  |  |
| Spouse/partner | 51 | (52.0) | 102 | (50.0) | 153 | (50.7) |
| Child | 46 | (46.9) | 91 | (44.6) | 137 | (45.4) |
| Friend | 0 | (0.0) | 1 | (0.5) | 1 | (0.3) |
| Other | 1 | (1.0) | 10 | (4.9) | 11 | (3.6) |
| Carer Living situation, n(%) |  |  |  |  |  |  |
| Live alone | 4 | (4.1) | 9 | (4.4) | 13 | (4.3) |
| Live with partner/spouse | 69 | (70.4) | 148 | (72.5) | 217 | (71.9) |
| Live with flat/housemates | 0 | (0.0) | 2 | (1.0) | 2 | (0.7) |
| Live with parent(s) | 13 | (13.3) | 14 | (6.9) | 27 | (8.9) |
| Live with children | 2 | (2.0) | 8 | (3.9) | 10 | (3.3) |
| Other | 10 | (10.2) | 23 | (11.3) | 33 | (10.9) |
| Carer Accommodation, n(%) |  |  |  |  |  |  |
| Council rented | 5 | (5.1) | 10 | (4.9) | 15 | (5.0) |
| Housing association rented | 4 | (4.1) | 6 | (2.9) | 10 | (3.3) |
| Private rented | 3 | (3.1) | 13 | (6.4) | 16 | (5.3) |
| Owner-occupied | 85 | (86.7) | 169 | (82.8) | 254 | (84.1) |
| Other | 1 | (1.0) | 6 | (2.9) | 7 | (2.3) |

**GCSE=General Certificate of Secondary Education. HNC=Higher National Certificate. HND=Higher National Diploma. NVQ=National Vocational Qualification.**

**Table 3:** Complete case mean cost of resource use at baseline 6-months and 12-months.

|  |  | Intervention |  | Control |  |
| --- | --- | --- | --- | --- | --- |
|  |  | N | mean (SD) (£) | N | mean (SD) (£) |
| Carer support services | Baseline | 204 | 253 (1061) | 98 | 98 (462) |
|  | 6 months | 141 | 320 (1229) | 66 | 370 (1170) |
|  | 12 months | 100 | 140 (574) | 59 | 271 (845) |
|  | total over 12 months | 90 | 520 (1549) | 53 | 510 (1307) |
| Community Health Services | Baseline | 204 | 211 (650) | 98 | 323 (738) |
|  | 6 months | 141 | 210 (770) | 66 | 274 (1102) |
|  | 12 months | 100 | 334 (999) | 59 | 200 (397) |
|  | total over 12 months | 90 | 620 (1924) | 53 | 504 (1621) |
| Emergency Care Services | Baseline | 204 | 141 (398) | 98 | 127 (280) |
|  | 6 months | 141 | 109 (263) | 66 | 53 (137) |
|  | 12 months | 100 | 125 (264) | 59 | 131 (292) |
|  | total at 12 | 90 | 252 (383) | 53 | 179 (339) |
| Hospital inpatient services | Baseline | 204 | 3567 (18590) | 98 | 1596 (8750) |
|  | 6 months | 141 | 4117 (21806) | 66 | 8658 (46493) |
|  | 12 months | 100 | 6154 (25117) | 59 | 8501 (38043) |
|  | total over 12 months | 90 | 11012 (36607) | 53 | 11180 (40507) |
| Day case treatment | Baseline | 204 | 105 (227) | 98 | 153 (449) |
|  | 6 months | 141 | 104 (198) | 66 | 71 (136) |
|  | 12 months | 100 | 57 (153) | 59 | 89 (245) |
|  | total over 12 months | 90 | 167 (286) | 53 | 165 (324) |
| Outpatient attendances | Baseline | 204 | 193 (456) | 98 | 213 (335) |
|  | 6 months | 141 | 195 (458) | 66 | 140 (220) |
|  | 12 months | 100 | 205 (328) | 59 | 230 (578) |
|  | total over 12 months | 90 | 439 (752) | 53 | 344 (675) |
| Home adaptations | Baseline | 204 | 2237 (2786) | 98 | 1957 (2712) |
|  | 6 months | 141 | 1946 (2665) | 66 | 1259 (2296) |
|  | 12 months | 100 | 1796 (2725) | 59 | 1367 (2595) |
|  | total over 12 months | 90 | 3650 (4602) | 53 | 2890 (4327) |
| State Funded Social care | Baseline | 204 | 61 (345) | 98 | 10 (91) |
|  | 6 months | 141 | 37 (260) | 66 | 0 (0) |
|  | 12 months | 100 | 15 (113) | 59 | 679 (2504) |
|  | total over 12 months | 90 | 33 (180) | 53 | 519 (2052) |
| Medication cost | Baseline | 204 | 143 (438) | 98 | 447 (2537) |
|  | 6 months | 141 | 43 (117) | 66 | 172 (889) |
|  | 12 months | 100 | 106 (590) | 59 | 73 (170) |
|  | total over 12 months | 90 | 152 (631) | 53 | 279 (1002) |
| State funded care home | 6 months | 141 | 219 (1283) | 66 | 488 (2386) |
|  | 12 months | 100 | 1060 (4714) | 59 | 187 (895) |
|  | total over 12 months | 90 | 1205 (4972) | 53 | 434 (2156) |
| **Total health and** | **Baseline** | **204** | **7040 (19920)** | **98** | **5054 (9672)** |
| personal social services * | **6 months** | **148** | **7360 (22625)** | **70** | **11089 (44972)** |
|  | **12 months** | **111** | **8536 (24916)** | **67** | **10906 (36402)** |
|  | **total at 12** | **101** | **16595 (37089)** | **61** | **24062 (60985)** |
| Social care private | Baseline | 204 | 381 (881) | 98 | 373 (908) |
|  | 6 months | 141 | 281 (1493) | 66 | 221 (847) |
|  | 12 months | 100 | 1209 (1645) | 59 | 978 (1486) |
|  | total at 12 | 90 | 1359 (1768) | 53 | 1180 (1684) |
| Social care unpaid | Baseline | 204 | 3142 (3475) | 98 | 2762 (2932) |
|  | 6 months | 141 | 1364 (1935) | 66 | 793 (1177) |
|  | 12 months | 100 | 1714 (2298) | 59 | 1131 (1656) |
|  | total at 12 | 90 | 3063 (3225) | 53 | 1787 (2064) |
| out-of-pocket payments | Baseline | 204 | 1975 (21897) | 98 | 651 (4071) |
|  | 6 months | 141 | 79 (581) | 66 | 6 (43) |
|  | 12 months | 100 | 108 (812) | 59 | 238 (1823) |
|  | total at 12 | 90 | 236 (1128) | 53 | 272 (1923) |
| **Total Wider Societal Costs\*** | **Baseline** | **204** | **5742 (21989)** | **98** | **4007 (5434)** |
|  | **6 months** | **149** | **1850 (2982)** | **70** | **1368 (2259)** |
|  | **12 months** | **111** | **2969 (3836)** | **67** | **2831 (5216)** |
|  | **total at 12** | **102** | **4702 (4884)** | **61** | **4037 (6091)** |

\* Includes a cost for people who died.

Table 4 shows imputed, adjusted total mean costs. The total mean imputed health and social care cost per participant at 12 months including the cost of training and delivering NIDUS-Family was £15,406 (SE 2,723) in the intervention arm compared to £23,867 (SE 7,450) in the control arm. Including the cost of the intervention, participants randomized to NIDUS-Family cost £8934 (37%) less (95% CI -£59,460 to £41,592) than the control group, for imputed costs and when adjusting for site, baseline costs and clustering for facilitator; this difference is not statistically significant. From a wider societal perspective, including out-of-pocket costs, time off work, and unpaid carer time, the mean cost per participant was £20,118 (SE 2746) for participants randomized to NIDUS-Family and £27,703 (SE 7436) for the control group, with a mean adjusted cost difference of -£8,176 (95% CI -£55,944 to £39,591).

**Table 4:** Mean costs and utility scores by study arm (Multiple Imputation data)

|  |  | Intervention<br>Mean (SE) |  | Control<br>Mean (SE) | Adjusted difference<br>(95% Confidence<br>Interval) |
| --- | --- | --- | --- | --- | --- |
| Health and Social Care Cost |  |  |  |  |  |
| Baseline | 204 | 7040 (1395) | 98 | 5054 (977) |  |
| 6 months | 204 | 6968 (1787) | 98 | 11439 (5313) | -4806 (-17188 to 7576) |
| 12 months | 204 | 8238 (1999) | 98 | 12,351 (5010) | -4289 (-21,515 to 12,937) |
| Total Health and Social care + intervention cost | 204 | 15,406 (2723) | 98 | 23,867 (7450) | -8934 (-59,460 to 41,592) |
| Wider societal Cost |  |  |  |  |  |
| Baseline | 204 | 5742 (1540) | 98 | 4007 (549) |  |
| 6 months | 204 | 1843 (210) | 98 | 1330 (227) | 495 (-84 to 1073) |
| 12 months | 204 | 3103 (308) | 98 | 3179 (549) | -136 (-1380 to 1108) |
| Total Wider Societal cost | 204 | 4954 (394) | 98 | 4517 (615) | -360 (-896 to 1617) |
| Overall total cost | 204 | 20,118 (2746) | 98 | 27702 (7436) | -8176 (-55,944 to 39,591) |
| DeMQoL-proxy utility scores & QALYs |  |  |  |  |  |
| Baseline utility | 203 | 0.794 (0.007) | 96 | 0.798 (0.009) | 0.008 (-0.025 to 0.041) |
| 6 months utility | 203 | 0.761 (0.016) | 96 | 0.754 (0.024) |  |
| 12 months utility | 203 | 0.748 (0.019) | 96 | 0.732 (0.027) |  |
| 12 months QALYs | 203 | 0.766 (0.012) | 96 | 0.761 (0.018) |  |
| CareQoL utility scores |  |  |  |  |  |
| Baseline CarerQoL | 203 | 77.36 (1.15) | 96 | 77.65 (1.88) |  |
| 6 months CarerQoL | 203 | 80.59 (1.20) | 96 | 77.69 (2.20) | 2.15 (-5.06 to 9.37) |
| 12 months CarerQoL | 203 | 80.62 (1.33) | 96 | 78.66 (1.78) | 1.50 (-3.96 to 6.96) |

Just over 10% (n=21) of the participants in the study moved to a care home (for respite-break or permanently) in the intervention group and 7% (n=7) in the control group, with 1 person in each group having more than one stay. There was no significant difference in the time spent in a care home between the two groups (adjusted mean difference: 0.117 weeks (95% CI -0.456 to 0.689).

### Utility scores

The unadjusted mean utility scores regarding client (DEMQOL-Proxy) and carer quality of life (CareQol) are reported in Table 4 (multiple imputation data), with complete case data reported in Table 3 (supplementary material). Complete data to calculate the DEMQOL-U index were 301 (99.7%) of participants at baseline, 205 (67.9%) participants at 6 months and 162 (53.6%) participants at 12 months. A zero was imputed for the 19 participants that died. There was a total of 164 (54.3%) participants with data available to calculate QALYs across the 12-months. For client quality of life, the mean imputed QALY at 12 months was 0.766 (95% CI: 0.743 to 0.789) for participants randomised to NIDUS-Family and 0.761 (95%CI: 0.726 to 0.796) for participants randomised to control, with a mean difference in QALYs of 0.008 (95%CI: - 0.025 to 0.041) adjusting for baseline utility, site and clustering for facilitator. Carers randomised to NIDUS-Family had a CareQoL tariff score 2.15 (95% CI -5.06 to 9.37) higher at 6-months and 1.50 higher (95% CI - 3.96 to 6.96) at 12-months compared to control, adjusting for baseline CareQoL, site and clustering for facilitator; these differences are not significant in the multiple imputation analysis (Table 4) (though attain significance in complete case analyses, Table 2S, supplemental material).

### Cost effectiveness analysis

The results of the 2-part bootstrap using the Multiple Imputation (MI) data and adjusting for baseline, site and facilitator clustering are reported in Figures 1a-1b for the health and personal social services perspective and Figures 1c-1d for the wider societal perspective. Across all analyses NIDUS-Family cost less for more QALYs compared to control but the difference was not statistically significant. At both a £20,000 and £30,000 decision threshold for a QALY gained from a client perspective there is an 89% probability that NIDUS-Family is cost-effective compared to usual care from a health and personal social services perspective and 87% probability that it is cost-effective from a wider societal perspective.

**Figure 1a:**
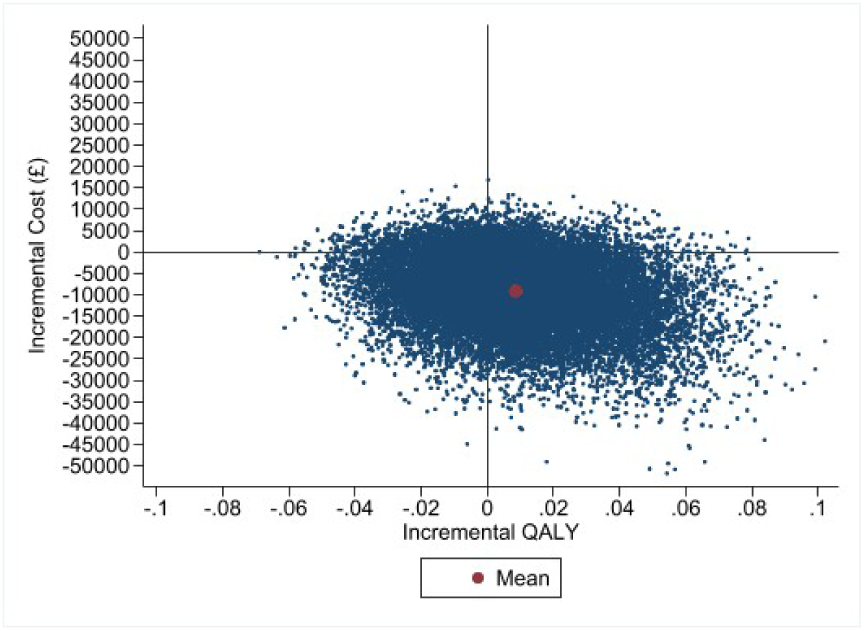
Cost effectiveness plane (NHS and personal social services perspective)

**Figure 1b:**
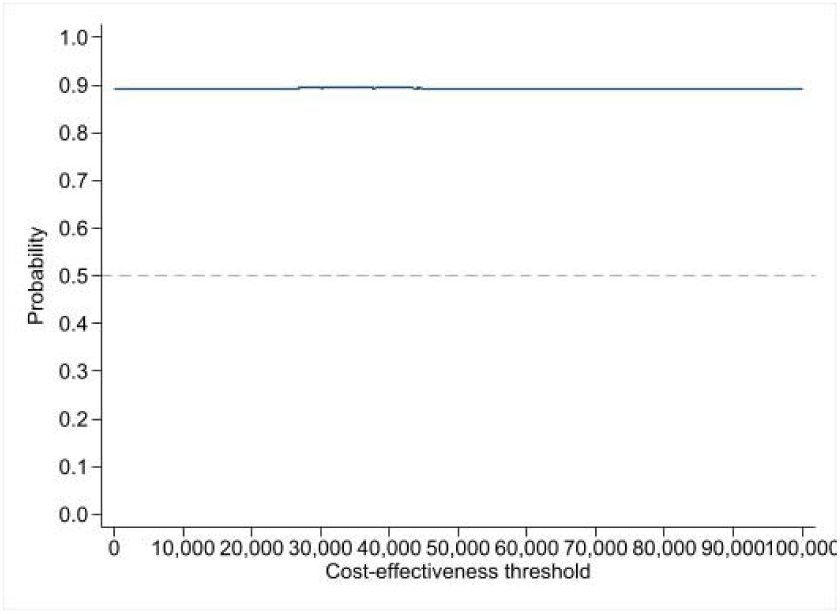
Cost effectiveness acceptability curve (NHS and personal social services perspective).

**Figure 1c:**
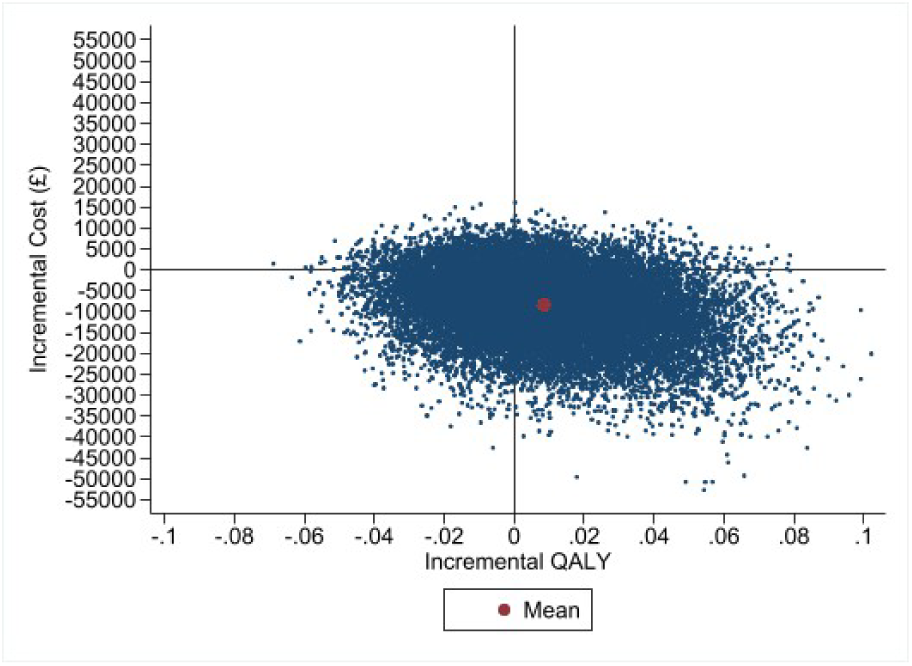
Cost effectiveness plane (Wider Societal perspective).

**Figure 1d:**
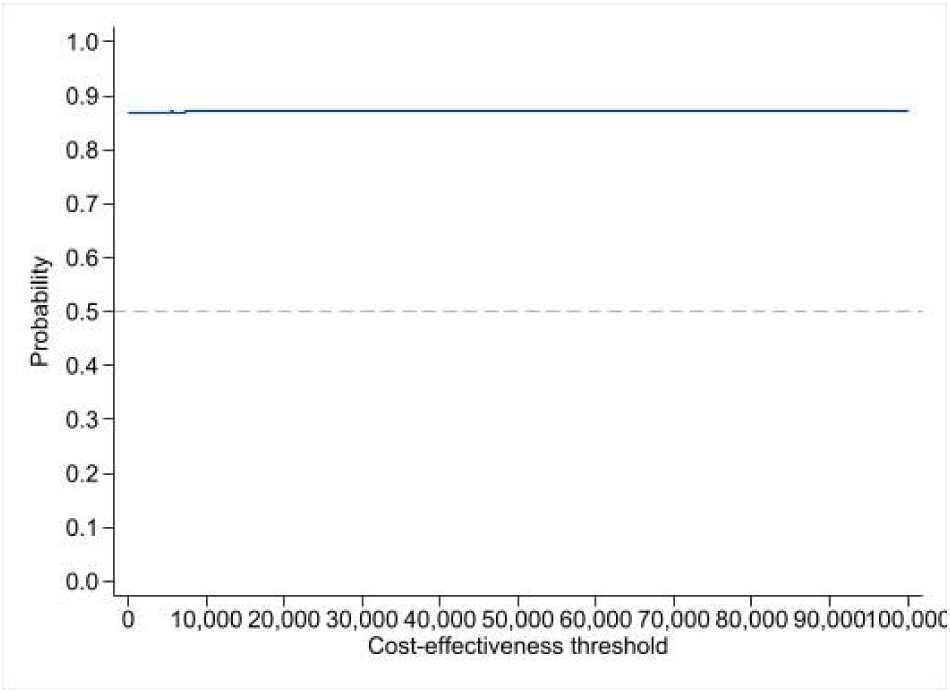
Cost effectiveness acceptability curve (Wider Societal perspective).

## Discussion

Our findings indicate a high probability that adding the NIDUS-Family intervention to treatment as usual is cost-effective, compared to treatment as usual over 12 months. Lower inpatient attendances and state-funded social care use contributed to intervention participants with dementia accruing £8934 (37%) lower costs from health and personal social services perspectives over a year than control arm participants, even after accounting for intervention costs. This cost difference was not statistically significant. The confidence intervals in the multiple imputation analysis are large because 12-month data was only available for 162 (53.6%) of participants, but even so there is an 89% probability that the intervention is cost-effective due to 88% of bootstrap iterations having a negative cost.

We identified a higher cost saving relative to usual care over a year than described for previous individually-delivered dementia care interventions in a recent review (6). In the review, the only comparable cost savings relative to usual care were associated with a group-based exercise intervention ($12,055 per dyad)(27). The flexibility of NIDUS-Family, to be delivered as an individual or dyadic intervention, remotely or in person, means it is widely scalable, with potential to reduce geographical inequalities in access to psychosocial interventions for people with dementia and their families (28). This flexibility, and the potential of NIDUS-family to reduce inpatient and social care costs through greater focus on prevention and wellbeing mean it has potential to reduce concerning care inequalities reported in dementia (3).

The cost of NIDUS-Family intervention was relatively low because facilitators were trained and supervised, but did not have formal clinical training, and it could be delivered remotely. A significant proportion of intervention costs were for group supervision and training provided by a senior clinician. If the intervention were delivered at scale, then economies of scale would reduce the supervision-associated cost per dyad. In the START (STrATegies for carers) intervention trial, which used a similar delivery model, client-related intervention costs were initially higher than usual care costs (10), but over six years, intervention arm cost savings increased, suggesting that benefits of psychosocial-based therapies that prevent distress accumulate over time (29). We have extended data collection up to two years, so will report later whether NIDUS-Family increased time living at home (without permanent care home move) over that period.

We conducted a range of analyses to evaluate the impact of missing data. As in all psychosocial treatment trials, we could not blind participants to allocation status. We are currently planning implementation studies and will explore how delivery in routine care compares with the trial setting, where it was delivered by a dedicated and well-resourced team (11).

NIDUS-Family is cost-effective, with a dominant intervention. It is the first personalised care and support intervention (13,14), to demonstrate effectiveness from the perspective of the quality of life of people with dementia deliverable by non-clinical facilitators and is thus potentially more scalable than other current personalised dementia care interventions. It was associated with a non-statistically significant client QALY gain and an 89% probability of cost-effectiveness. The care approach used by NIDUS-Family aligns with aspirations of the NHS Long Term Workforce Plan, to innovate and grow the healthcare workforce (30). Routine provision of NIDUS-Family within post-diagnostic services to all people diagnosed with dementia with a family carer could be transformative; we are currently conducting an implementation study to explore how it works in routine practice.

### Research in Context Panel

#### Evidence before this study

A systematic review (PROSPERO CRD42021252999) of economic evaluations of non-pharmacological interventions for dementia or mild cognitive impairment searched the following databases: Academic Search Premier, MEDLINE, Web of Science, EMBASE, Google Scholar, Cumulative Index to Nursing and Allied Health Literature (CINAHL), PsycInfo, Psychology and Behavioural Sciences Collection, PsycArticles, Cochrane Database of Systematic Reviews, Business Source Premier and Regional Business News; up to May 2023. It found the strongest evidence of cost-effectiveness was for Maintenance Cognitive Stimulation Therapy (MCST). Case management, occupational therapy and dementia care management also showed good evidence of cost-effectiveness. The authors concluded that more economic evidence on the cost-effectiveness of specific dementia care interventions could improve local and national decision makers’ confidence to promote future cost-effective dementia interventions (6). We updated the search and found one more relevant publication, which calculated the costs of scaling up START, a manualized intervention that reduced and prevented anxiety and depression symptoms in family carers of people with dementia, and had a high probability of cost-effectiveness for family carers wellbeing. The authors estimated that scaling up the START intervention to eligible carers would cost £9.4 million in 2020, but these costs would lead to annual savings of £68 million, and total annual quality-adjusted life year (QALY) gains of 1247 (31). We previously reported that the NIDUS-Family intervention was effective in increasing attainment of dyadic goals. It is, to our knowledge, the first intervention to improve goal attainment in people living with dementia that is scalable and can be delivered by people without clinical training; and that can be delivered remotely (11).

#### Added value of this study

NIDUS-Family is cost-effective, from health and social care and societal perspectives, as well as clinically effective and should be part of routine dementia care. It is the first personalised care and support intervention (12,27), to demonstrate effectiveness from the perspective of the quality of life of people with dementia deliverable by non-clinical facilitators. It is thus potentially more scalable than other current personalised dementia care interventions. It was associated with a non-statistically significant client QALY gain and an 89% probability of cost-effectiveness.

#### Implications of all the available evidence

NIDUS-Family is inexpensive, scalable, and inclusive. The few non-pharmacological interventions demonstrating effectiveness in RCTs involving people living with dementia have been planned around personal goals, and NIDUS-Family is to our knowledge the first evidence-based, manualised intervention that can enable such care that has been shown to be cost-effective. We recommend that post-diagnostic services routinely provide goal-focused, structured, manualised support to all people diagnosed with dementia with a regular carer.

## Data Availability

All data produced in the present study are available upon reasonable request to the authors

## Authors’ Contributors

AI and RH designed the analysis, and RH reviewed it as senior health economist. AI, RH and CC drafted the paper. CC was Chief Investigator of the NIDUS study. At least two authors accessed and verified the data (AI, RH). JBu managed the trial. SB chaired the independent steering committee. KR provided expert guidance and training on use of GAS. PR and CC supervised the intervention delivery. All authors had full access to all the data in the study and had final responsibility for the decision to submit for publication.

## Data sharing

Data collected for the study, including the statistical analysis plan, deidentified participant data and a data dictionary defining each field in the set, will be made available to others on receipt by Priment CTU of a reasonable request, at any date after publication of this paper. All requests will be reviewed by Priment CTU in line with Priment CTU guidance on sharing data and anonymising data. This process is to ensure that the request is reasonable and the data set is suitably anonymised. The study protocol is available open access. Intervention materials are available without cost, subject to a CC BY-NC-ND license held by Claudia Cooper, Chief Investigator.

## Acknowledgments

This work was supported by the Alzheimer’s Society (Centre of Excellence grant 330). We thank the NIDUS participants and PPI group. IL’s time is supported in part by the UK National Institute for Health and Care Research (NIHR) Applied Research Collaboration South West Peninsula (Grant Reference Number: NIHR200167).

SB declares grants from National Institute for Health and Care Research (NIHR), Economic and Social Research Council, Engineering and Physical Science Research Council, Canadian Institute for Health Research, the Alzheimer’s Association, the Alzheimer’s Society, Health Education England. He has held the following positions: Non-Executive Director Somerset NHS Foundation Trust, Trustee of the Alzheimer’s Society, Executive Dean of the University of Plymouth, and Pro-Vice Chancellor of the University of Nottingham. He has acted as a consultant and in an educational role for Lilly and Lundbeck. Claudia Cooper, Iain Lang, Rachael Hunter and Sube Banerjee are supported by the National Institute for Health and Care Research (NIHR) Dementia and Neurodegeneration Policy Research Unit (NIHR206110). Claudia Cooper is supported by an NIHR Senior Investigator award (NIHR205009). The views expressed are those of the author(s) and not necessarily those of the NIHR, the NHS or the Department of Health and Social Care.

