## Supplemental material for "Cost-utility of a new psychosocial goal-setting and manualised support intervention for Independence in Dementia (NIDUS-Family) versus goal-setting and routine care: economic evaluation embedded within a randomised controlled trial"

**Supplementary Material**

Supplementary Table 1: Unit costs

| **Variable name** | **Unit Cost £ (2022/23)** | **Unit** | **Source** | **Notes/Assumptions** |
| --- | --- | --- | --- | --- |
| GP Consultation at surgery | 42 | per surgery consultation | PSSRU 2022.  Table 9.4.2 pg 70 | GP consultation multiplied by surgery:clinic ratio.  This cost is based on GP consultation of an average 9.22 minutes per consultation |
| GP telephone consultation | 17 | per e-consultation | PSSRU 2022. Table 9.5.1, pg 72 | GP consultation multiplied by surgery:phone ratio. |
| GP at home consultation | 70 | per home consultation |  | GP consultation multiplied by surgery:home ratio |
| Nurse practice at clinic | 13 | per consultation | PSSRU 2022.  Table 9.3.1 pg 68 | Hourly wage of a practice nurse multiplied by surgery appointment time from PSSRU 2015. |
| Nurse practice by telephone | 5 | per consultation | PSSRU 2022.  Table 9.3.1 pg 68 | Hourly wage of a practice nurse multiplied by phone appointment time from PSSRU 2015. |
| Nurse practice at home | 22 | per consultation | PSSRU 2022.  Table 9.3.1 pg 68 | Hourly wage of a practice nurse multiplied by home appointment time from PSSRU 2015 |
| Admiral nurse at clinic | 15.75 | per consultation | PSSRU 2022.  Table 9.2.1 pg 66 | Admiral nurse is specialist nurse for dementia at Afc band 6. Hourly wage of band 6 nurse multiplied by surgery nurse appoinment time from PSSRU 2015 |
| Admiral nurse by telephone | 6.3 | per consutation | PSSRU 2022.  Table 9.2.1 pg 66 | Admiral nurse is specialist nurse for dementia at Afc band 6. Hourly wage of band 6 nurse multiplied by phone nurse appoinment time from PSSRU 2015 |
| Admiral nurse at home | 26 | per consutation | PSSRU 2022.  Table 9.2.1 pg 66 | Admiral nurse is specialist nurse for dementia at Afc band 6. Hourly wage of band 6 nurse multiplied by home nurse appoinment time from PSSRU 2015 |
| Community/district nurse at clinic | 14.41 | per consultation | PSSRU 2015.  Table 10.1 pg 169 | Community/District nurse hourly wage multiplied by surgery nurse appoinment time from PSSRU 2015 and uprated to 2022. |
| Community/district nurse by telephone | 5.77 | per consultation | PSSRU 2015.  Table 10.1 pg 169 | Community/District nurse hourly wage multiplied by phone nurse appoinment time from PSSRU 2015 and uprated to 2022. |
| Community/district nurse at home | 24 | consultation | PSSRU 2015.  Table 10.1 pg 169 | Community/District nurse hourly wage multiplied by home nurse appoinment time from PSSRU 2015 and uprated to 2022. |
| Memory service doctor-at clinic | 123 | per consultation | PSSRU 2017. Table 1.7, pg 39 | No information available on ratio of contact times so applied the practice nurse contact time from PSSRU 2015. Assumed hourly consultation wage thus divided the team cost by 60 minutes before applying the practice nurse ratio for surgery appointment time.Uprated to 2022. |
| Memory service doctor - by telphone | 49 | per consultation | PSSRU 2017. Table 1.7, pg 39 | No information available on ratio of contact times so applied the practice nurse contact time from PSSRU 2015. Assumed hourly consultation wage thus divided the team cost by 60 minutes before applying the practice nurse ratio for phone appointment time.Uprated to 2022 |
| Memory service doctor - at home | 205 | per consultation | PSSRU 2017. Table 1.7, pg 39 | No information available on ratio of contact times so applied the practice nurse contact time from PSSRU 2015. Assumed hourly consultation wage thus divided the team cost by 60 minutes before applying the practice nurse ratio for home appointment time.Uprated to 2022. |
| Memory service psychologist -at clinic (hospital based) | 16 | per consultation | PSSRU 2022. Table 11.1.2, pg 95 | No information available on ratio of contact times so applied the practice nurse contact time from PSSRU 2015. Hourly consultation cost divided by 60 minutes before applying the practice nurse ratio for surgery appointment time. |
| Memory service psychologist -by telephone (hospital based) | £6 | per consultation | PSSRU 2022. Table 11.1.2, pg 95 | No information available on ratio of contact times so applied the practice nurse contact time from PSSRU 2015. Hourly consultation cost divided by 60 minutes before applying the practice nurse ratio for phone appointment time. |
| Memory service psychologist -at home (hospital based) | 27 | per consultation | PSSRU 2022. Table 11.1.2, pg 95 | No information available on ratio of contact times so applied the practice nurse contact time from PSSRU 2015. Hourly consultation cost divided by 60 minutes before applying the practice nurse ratio for home appointment time. |
| Memory service nurse - at clinic | 12 | per consultation | PSSRU 2022. Table 11.2.2, pg 97 | No information available on ratio of contact times so applied the practice nurse contact time from PSSRU 2015. Hourly consultation cost divided by 60 minutes before applying the practice nurse ratio for surgery appointment time. |
| Memory service nurse - by telephone | 5 | per consultation | PSSRU 2022. Table 11.2.2, pg 97 | No information available on ratio of contact times so applied the practice nurse contact time from PSSRU 2015. Hourly consultation cost divided by 60 minutes before applying the practice nurse ratio for phone appointment time. |
| Memory service nurse - at home | 20 | per consultation | PSSRU 2022. Table 11.2.2, pg 97 | No information available on ratio of contact times so applied the practice nurse contact time from PSSRU 2015. Hourly consultation cost divided by 60 minutes before applying the practice nurse ratio for home appointment time. |
| Memory service assistant practitioner - at clinic | 9 | per consultation | PSSRU 2022. Table 8.2.1, pg 60 | Assistant practioner are paid on NHS Afc band 4 rate.No information available on ratio of contact times so applied the practice nurse contact time from PSSRU 2015. Hourly consultation cost divided by 60 minutes before applying the practice nurse ratio for surgery appointment time. |
| Memory service assistant practitioner - by telephone | 4 | per consultation | PSSRU 2022. Table 8.2.1, pg 60 | Assistant practioner are paid on NHS Afc band 4 rate.No information available on ratio of contact times so applied the practice nurse contact time from PSSRU 2015. Hourly consultation cost divided by 60 minutes before applying the practice nurse ratio for phone appointment time. |
| Memory service assistant practitioner - at home | 15 | consultation | PSSRU 2022. Table 8.2.1, pg 60 | Assistant practioner are paid on NHS Afc band 4 rate.No information available on ratio of contact times so applied the practice nurse contact time from PSSRU 2015. Hourly consultation cost divided by 60 minutes before applying the practice nurse ratio for home appointment time. |
| Community Mental Health Team (CMHT) doctor - at clinic | 12 | consultation | PSSRU 2015. Table 12.1, pg 199 | No information available on ratio of contact times so applied the practice nurse contact time from PSSRU 2015. Hourly consultation cost divided by 60 minutes before applying the practice nurse ratio for surgery appointment time. The unit cost is taken from the unit cost for the team as per PSSRU 2015 and inflated using 2022 inflation index.Uprated to 2022 |
| Community Mental Health Team (CMHT) doctor - by telephone | 5 | per consutation | PSSRU 2015. Table 12.1, pg 199 | No information available on ratio of contact times so applied the practice nurse contact time from PSSRU 2015. Hourly consultation cost divided by 60 minutes before applying the practice nurse ratio for phone appointment time. The unit cost is taken from the unit cost for the team as per PSSRU 2015 and inflated using 2022 inflation index. Uprated to 2022 |
| Community Mental Health Team (CMHT) doctor - at home | 20 | per consultation | PSSRU 2015. Table 12.1, pg 199 | No information available on ratio of contact times so applied the practice nurse contact time from PSSRU 2015. Hourly consultation cost divided by 60 minutes before applying the practice nurse ratio for home appointment time. The unit cost is taken from the unit cost for the team as per PSSRU 2015 and inflated using 2022 inflation index. Uprated to 2022 |
| CMHT nurse - at clinic | 12 | per consultation | PSSRU 2015. Table 12.1, pg 199 | No information available on ratio of contact times so applied the practice nurse contact time from PSSRU 2015. Hourly consultation cost divided by 60 minutes before applying the practice nurse ratio for surgery appointment time. The unit cost is taken from the unit cost for the team as per PSSRU 2015 and inflated using 2022 inflation index. Uprated to 2022 |
| CMHT nurse - by phone | 5 | per consultation | PSSRU 2015. Table 12.1, pg 199 | No information available on ratio of contact times so applied the practice nurse contact time from PSSRU 2015. Hourly consultation cost divided by 60 minutes before applying the practice nurse ratio for phone appointment time. The unit cost is taken from the unit cost for the team as per PSSRU 2015 and inflated using 2022 inflation index. Uprated to 2022 |
| CMHT nurse - at home | 20 | per consultation | PSSRU 2015. Table 12.1, pg 199 | No information available on ratio of contact times so applied the practice nurse contact time from PSSRU 2015. Hourly consultation cost divided by 60 minutes before applying the practice nurse ratio for home appointment time. The unit cost is taken from the unit cost for the team as per PSSRU 2015 and inflated using 2022 inflation index. Uprated to 2022 |
| CMHT psychologist - at clinic | 17 | per consultation | PSSRU 2022. Table 8.2.1, pg 60 | Used community based clinical psychologist band 7.No information available on ratio of contact times so applied the practice nurse contact time from PSSRU 2015. Hourly consultation cost divided by 60 minutes before applying the practice nurse ratio for surgery appointment time. |
| CMHT psychologist - by phone/video | 7 | per consultatation | PSSRU 2022. Table 8.2.1, pg 60 | Used community based clinical psychologist band 7.No information available on ratio of contact times so applied the practice nurse contact time from PSSRU 2015. Hourly consultation cost divided by 60 minutes before applying the practice nurse ratio for phone appointment time. |
| CMHT psychologist - at home | 28 | per consultation | PSSRU 2022. Table 8.2.1, pg 60 | Used community based clinical psychologist band 7.No information available on ratio of contact times so applied the practice nurse contact time from PSSRU 2015. Hourly consultation cost divided by 60 minutes before applying the practice nurse ratio for home appointment time. |
| CMHT assistant practitioner - at clinic | 9 | per consultation | PSSRU 2022.  Table 8.2.1 pg 60 | CMHT assistant psychologist is paid at NHS Afc band 4.No information available on ratio of contact times so applied the practice nurse contact time from PSSRU 2015. Hourly consultation cost divided by 60 minutes before applying the practice nurse ratio for surgery appointment time. |
| CMHT assistant practitioner - by phone/video | 4 | per consultation | PSSRU 2022.  Table 8.2.1 pg 60 | CMHT assistant psychologist is paid at NHS Afc band 4.No information available on ratio of contact times so applied the practice nurse contact time from PSSRU 2015. Hourly consultation cost divided by 60 minutes before applying the practice nurse ratio for phone appointment time. |
| CMHT assistant practitioner - at home | 15 | per consultation | PSSRU 2022.  Table 8.2.1 pg 60 | CMHT assistant psychologist is paid at NHS Afc band 4.No information available on ratio of contact times so applied the practice nurse contact time from PSSRU 2015. Hourly consultation cost divided by 60 minutes before applying the practice nurse ratio for home appointment time. |
| Occupational therapist(OT) - at clinic | 14 | per consultation | PSSRU 2022. Table 8.2.1, pg 60 | Community based OT band 6.No information available on ratio of contact times so applied the practice nurse contact time from PSSRU 2015. Hourly consultation cost divided by 60 minutes before applying the practice nurse ratio for surgery appointment time. |
| Occupational therapist - by phone/video | 6 | per consultation | PSSRU 2022. Table 8.2.1, pg 60 | Community based OT band 6.No information available on ratio of contact times so applied the practice nurse contact time from PSSRU 2015. Hourly consultation cost divided by 60 minutes before applying the practice nurse ratio for phone appointment time. |
| Occupational therapist - at home | 23 | per consultation | PSSRU 2022. Table 8.2.1, pg 60 | Community based OT band 6.No information available on ratio of contact times so applied the practice nurse contact time from PSSRU 2015. Hourly consultation cost divided by 60 minutes before applying the practice nurse ratio for home appointment time. |
| Physiotherapist(PT) - at clinic | 14 | consultation | PSSRU 2022. Table 8.2.1, pg 60 | Community based PT band 6.No information available on ratio of contact times so applied the practice nurse contact time from PSSRU 2015. Hourly consultation cost divided by 60 minutes before applying the practice nurse ratio for surgery appointment time. |
| Physiotherapist - by phone/video | 6 | per consultation | PSSRU 2022. Table 8.2.1, pg 60 | Community based PT band 6.No information available on ratio of contact times so applied the practice nurse contact time from PSSRU 2015. Hourly consultation cost divided by 60 minutes before applying the practice nurse ratio for phone appointment time. |
| Physiotherapist - at home | 23 | per consultation | PSSRU 2022. Table 8.2.1, pg 60 | Community based PT band 6.No information available on ratio of contact times so applied the practice nurse contact time from PSSRU 2015. Hourly consultation cost divided by 60 minutes before applying the practice nurse ratio for home appointment time. |
| Specialist nurse - at clinic | 13 | per consultation | PSSRU 2022. Table 11.2.2, pg 97 | Nurse specialist hospital based at band 6. No information avaialable on ratio of contact times so applied the practice nurse contact time from PSSRU 2015.  Hourly consultation cost divided by 60 minutes before applying the practice nurse ratio for surgery appointment time. |
| Specialist nurse - by phone/video | 5 | consultation | PSSRU 2022. Table 11.2.2, pg 97 | Nurse specialist hospital based at band 6. No information avaialable on ratio of contact times so applied the practice nurse contact time from PSSRU 2015.  Hourly consultation cost divided by 60 minutes before applying the practice nurse ratio for phone appointment time. |
| Specialist nurse - at home | 22 | per consultation | PSSRU 2022. Table 11.2.2, pg 97 | Nurse specialist hospital based at band 6. No information avaialable on ratio of contact times so applied the practice nurse contact time from PSSRU 2015.  Hourly consultation cost divided by 60 minutes before applying the practice nurse ratio for home appointment time. |
| Other community Practitioner  - at clinic | 11 | per consultation | PSSRU 2022. Table 8.2.1, pg 60 | Comm. Practioner level varies, assumed band 5 comm nurse  level PSSRU 2022. No ifnormation available on ratio of contact times so applied the hourly cost divided by 60 minutes before applying the practice nurse ratio for surgery appointment time. |
| Other community Practitioner - by phone/video | 4 | per consultation | PSSRU 2022. Table 8.2.1, pg 60 | Comm. Practioner level varies, assumed band 5 comm nurse  level PSSRU 2022. No ifnormation available on ratio of contact times so applied the hourly cost divided by 60 minutes before applying the practice nurse ratio for phone appointment time. |
| Other community Practitioner - at home | 18 | per consultation | PSSRU 2022. Table 8.2.1, pg 60 | Comm. Practioner level varies, assumed band 5 comm nurse  level PSSRU 2022. No ifnormation available on ratio of contact times so applied the hourly cost divided by 60 minutes before applying the practice nurse ratio for home appointment time. |
| Carer support programme - at clinic | 13 | per consultation | PSSRU 2022  table 10.1.1 pg 81 | Assumed social worker level for carer support programme. No ifnormation available on ratio of contact times so applied the hourly cost divided by 60 minutes before applying the practice nurse ratio for surgery appointment time. |
| Carer support programme - by phone/video | 5 | per consultation | PSSRU 2022 Table 10.1.1 pg 81 | Assumed social worker level for carer support programme. No ifnormation available on ratio of contact times so applied the hourly cost divided by 60 minutes before applying the practice nurse ratio for phone appointment time. |
| Carer support programme - at home | 21 | per consultraion | PSSRU 2022 Table 10.1.1 pg 81 | Assumed social worker level for carer support programme. No ifnormation available on ratio of contact times so applied the hourly cost divided by 60 minutes before applying the practice nurse ratio for home appointment time. |
| Other services - at clinic | 13 | per visit | PSSRU 2022 ,  table 4.3.1, pg 31 | A variety of other services exist so assumed other services for used social worker. No contact information so applied the practice nurse ratio for surgery apppointment |
| Other services - by phone/video | 5 | per visit | PSSRU 2022 ,  table 4.3.1, pg 31 | A variety of other services exist so assumed other services for used social worker. No contact information so applied the practice nurse ratio for phone apppointment |
| Other services - at home | 21 | per visit | PSSRU 2022 ,  Table 4.3.1, pg 31 | A variety of other services exist so assumed other services for used social worker. No contact information so applied the practice nurse ratio for home appointment. |
| NHS direct or "call 111" | 12 | per call | Turner J, et al(2021).  Impact of NHS 111 Online  on the NHS 111 telephone  service and urgent  care system: a  mixed-methods  study. Health Serv Deliv Res. | No information on cost of NHS call cost available on PSSRU, so used information on the Turner J et al(2021) study.Uprated to 2022. |
| Emergency call(999) | 12 | per call | Turner J, et al(2021).  Impact of NHS 111 Online  on the NHS 111 telephone  service and urgent  care system: a  mixed-methods  study. Health Serv Deliv Res. | Assumed cost as in NHS 111 call from Turner J , et al(2021) Uprated to 2022. |
| Paramedic only | 237 | per service | NHS referecence cost- EC services. 2021/2022 |  |
| Paramedic and ambulance to hospital | 242 | per service | NHS referecence cost- EC services. 2021/2022 |  |
| A&E attendance without ambulance | 154 | per service | NHS referecence cost- EC services. 2021/2022 |  |
| non-elective (emergency) inpatient stay for physical health | 985 | per service | NHS referecence cost- EC services. 2021/2022 |  |
| Elective inpatient stay for physical health | 5845 | per service | Average national cost  2021/2022 (NHS reference cost) |  |
| Inpatient hospital stay for mental health | 342 | per service | NHS reference cost  MH services 2021/2022 |  |
| General medical outpatient appointment | 235 | per service | NHS referecence cost- EC services. 2021/2022 |  |
| Day patient procedures/test | 203 | per service | NHS referecence cost- EC services. 2021/2022 |  |
| Psychiatric outpatient appointment | 342 | per service | NHS referecence cost- EC services. 2021/2022 |  |

**Figure 1: Consort** **diagram**

* Numbers are those providing **any** data at 6 months. For 52 (routine care = 19, NIDUS = 33) this was for the 6 month GAS outcome only.

**GAS was scored and analysed in the first follow-up post death. That is, deaths between 6 and 12 months are scored and analysed at 12 months (6 in routine care, 4 in NIDUS intervention)

Plwd = person living with dementia only minor - guidelines by people with dementia say we should avoid acronyms like that (PlWd)

Randomised

(n= 302)

Allocated to Routine Care

(n = 98)

Allocated to NIDUS intervention

(n = 204)

Received intervention (n=192)

Did not receive intervention (n=12)

Followed up at 6 months (n=83)*

CSRI complete (n=63)

Followed up at 6 months (n=169)*

No response at 6 months (n=6)

Loss to follow up (n=29):

Withdrawn (n=13)

No longer contactable (n=9)

Plwd died (n=6)

Carer died (n=1)

Loss to follow up (n=11):

Withdrawn (n=3)

No longer contactable (n=5)

Plwd died (n=3)

Number approached and assessed for eligibility (n=1,083)

(n=685)

Excluded (n=781):

Not eligible (n=581)

Declined to participate (n=200)

12 months primary outcome analysed (n=84)**

12 months primary outcome analysed (n=163)**

Loss to follow up (n=17):

Withdrawn (n=2)

No longer contactable (n=10)

Plwd died (n=5)

Loss to follow up (n=9):

Withdrawn (n=3)

Plwd died (n=6)

**Supplementary Table 2: Resource use descriptive statistics – the number of participants that used the service and the mean for those that used the service.**

|  | Intervention | | | Control | | |
| --- | --- | --- | --- | --- | --- | --- |
|  | Total N | n (%) | mean (SD) | Total N | n (%) | mean (SD) |
| **Proportion of carers in paid employment** |  |  |  |  |  |  |
| baseline | 204 | 91 (45%) |  | 98 | 41 (42%) |  |
| 6 months | 141 | 49 (35%) |  | 65 | 24 (37%) |  |
| 12 months | 100 | 41 (41%) |  | 59 | 22 (37%) |  |
| Days off work |  |  |  |  |  |  |
| baseline | 91 | 40 (44%) | 8 (12) | 40 | 16 (40%) | 7 (6) |
| 6 months | 47 | 20 (43%) | 10 (22) | 24 | 9 (38%) | 16 (15) |
| 12 months | 41 | 21 (51%) | 8 (11) | 22 | 9 (41%) | 26 (47) |
| 12 months total | 32 | 21 (66%) | 12 (24) | 18 | 9 (50%) | 29 (53) |
| **Carer services** |  |  |  |  |  |  |
| Admiral Nurse |  |  |  |  |  |  |
| baseline | 204 | 12 (6%) | 4 (4) | 98 | 4 (4%) | 2 (1) |
| 6 months | 141 | 16 (11%) | 3 (3) | 66 | 2 (3%) | 3 (3) |
| 12 months | 100 | 8 (8%) | 3 (2) | 59 | 1 (2%) | 6 (0) |
| 12 months total | 90 | 13 (14%) | 3 (2) | 53 | 2 (4%) | 6 (7) |
| Other carer services |  |  |  |  |  |  |
| baseline | 204 | 49 (24%) | 53 (93) | 98 | 27 (28%) | 21 (39) |
| 6 months | 141 | 39 (28%) | 59 (100) | 66 | 17 (26%) | 71 (93) |
| 12 months | 100 | 25 (25%) | 41 (73) | 59 | 20 (34%) | 42 (61) |
| 12 months total | 90 | 40 (44%) | 68 (108) | 53 | 22 (42%) | 64 (85) |
| **Paid and Unpaid Carer Time** |  |  |  |  |  |  |
| State funded -hours |  |  |  |  |  |  |
| baseline | 204 | 16 (8%) | 34 (44) | 98 | 2 (2%) | 20 (27) |
| 6 months | 141 | 4 (3%) | 57 (42) | 66 | 0 (0%) | 0 (0) |
| 12 months | 100 | 2 (2%) | 33 (18) | 59 | 5 (8%) | 348 (182) |
| 12 months total | 90 | 3 (3%) | 43 (4) | 53 | 4 (8%) | 299 (166) |
| Private -hours |  |  |  |  |  |  |
| baseline | 204 | 80 (39%) | 85 (116) | 98 | 36 (37%) | 84 (121) |
| 6 months | 141 | 43 (30%) | 86 (237) | 66 | 15 (23%) | 91 (146) |
| 12 months | 100 | 26 (26%) | 35 (66) | 59 | 12 (20%) | 79 (155) |
| 12 months total | 90 | 36 (40%) | 61 (107) | 53 | 19 (36%) | 122 (166) |
| Unpaid care -hours |  |  |  |  |  |  |
| baseline | 204 | 182 (89%) | 153 (152) | 98 | 93 (95%) | 127 (128) |
| 6 months | 141 | 110 (78%) | 76 (88) | 66 | 53 (80%) | 43 (54) |
| 12 months | 100 | 78 (78%) | 96 (104) | 59 | 47 (80%) | 62 (76) |
| 12 months total | 90 | 87 (97%) | 138 (140) | 53 | 49 (92%) | 84 (90) |
| **Community Health Care Services** |  |  |  |  |  |  |
| GP Contacts |  |  |  |  |  |  |
| baseline | 204 | 136 (67%) | 3 (4) | 98 | 73 (74%) | 9 (44) |
| 6 months | 141 | 95 (67%) | 3 (3) | 66 | 43 (65%) | 3 (3) |
| 12 months | 100 | 72 (72%) | 4 (4) | 59 | 42 (71%) | 4 (4) |
| 12 months total | 90 | 78 (87%) | 7 (6) | 53 | 47 (89%) | 5 (5) |
| Community Nurses |  |  |  |  |  |  |
| baseline | 204 | 96 (47%) | 7 (37) | 98 | 44 (45%) | 7 (19) |
| 6 months | 141 | 62 (44%) | 9 (46) | 66 | 24 (36%) | 19 (74) |
| 12 months | 100 | 56 (56%) | 11 (49) | 59 | 27 (46%) | 8 (19) |
| 12 months total | 90 | 66 (73%) | 16 (89) | 53 | 32 (60%) | 20 (81) |
| Memory Services |  |  |  |  |  |  |
| baseline | 204 | 102 (50%) | 2 (2) | 98 | 55 (56%) | 3 (3) |
| 6 months | 141 | 38 (27%) | 3 (6) | 66 | 22 (33%) | 3 (2) |
| 12 months | 100 | 28 (28%) | 5 (15) | 59 | 17 (29%) | 3 (3) |
| 12 months total | 90 | 38 (42%) | 6 (14) | 53 | 24 (45%) | 3 (3) |
| Community mental health teams |  |  |  |  |  |  |
| baseline | 204 | 14 (7%) | 4 (6) | 98 | 6 (6%) | 3 (1) |
| 6 months | 141 | 9 (6%) | 4 (5) | 66 | 2 (3%) | 2 (1) |
| 12 months | 100 | 9 (9%) | 3 (3) | 59 | 3 (5%) | 2 (2) |
| 12 months total | 90 | 17 (19%) | 4 (6) | 53 | 5 (9%) | 2 (1) |
| Other community services |  |  |  |  |  |  |
| baseline | 204 | 66 (32%) | 4 (8) | 98 | 35 (36%) | 8 (13) |
| 6 months | 141 | 49 (35%) | 3 (4) | 66 | 17 (26%) | 3 (3) |
| 12 months | 100 | 35 (35%) | 5 (6) | 59 | 26 (44%) | 4 (3) |
| 12 months total | 90 | 48 (53%) | 6 (6) | 53 | 29 (55%) | 5 (4) |
| **Secondary Care** |  |  |  |  |  |  |
| A&E attendances |  |  |  |  |  |  |
| baseline | 204 | 60 (29%) | 3 (2) | 98 | 29 (30%) | 3 (2) |
| 6 months | 141 | 44 (31%) | 2 (2) | 66 | 14 (21%) | 3 (3) |
| 12 months | 100 | 32 (32%) | 2 (1) | 59 | 19 (32%) | 2 (1) |
| 12 months total | 90 | 50 (56%) | 3 (2) | 53 | 20 (38%) | 3 (2) |
| Unplanned admissions - nights |  |  |  |  |  |  |
| baseline | 204 | 21 (10%) | 8 (9) | 98 | 9 (9%) | 5 (4) |
| 6 months | 141 | 14 (10%) | 9 (11) | 66 | 8 (12%) | 15 (24) |
| 12 months | 100 | 11 (11%) | 11 (11) | 59 | 8 (14%) | 13 (16) |
| 12 months total | 90 | 18 (20%) | 12 (13) | 53 | 10 (19%) | 12 (14) |
| Planned admissions - nights |  |  |  |  |  |  |
| baseline | 204 | 2 (1%) | 10 (6) | 98 | 0 (0%) | 0 (0) |
| 6 months | 141 | 1 (1%) | 3 (0) | 66 | 1 (2%) | 5 (0) |
| Mental Health Admissions - nights |  |  |  |  |  |  |
| baseline | 204 | 0 (0%) | 0 (0) | 98 | 1 (1%) | 1 (0) |
| 12 months | 100 | 2 (2%) | 33 (33) | 59 | 1 (2%) | 91 (0) |
| General Medical Outpatient |  |  |  |  |  |  |
| baseline | 204 | 66 (32%) | 2 (3) | 98 | 35 (36%) | 2 (2) |
| 6 months | 141 | 43 (30%) | 2 (2) | 66 | 23 (35%) | 2 (1) |
| 12 months | 100 | 37 (37%) | 2 (1) | 59 | 16 (27%) | 3 (3) |
| 12 months total | 90 | 50 (56%) | 3 (3) | 53 | 21 (40%) | 3 (3) |
| Psychiatric Outpatient |  |  |  |  |  |  |
| baseline | 204 | 2 (1%) | 1 (0) | 98 | 2 (2%) | 2 (1) |
| 6 months | 141 | 2 (1%) | 3 (3) | 66 | 1 (2%) | 1 (0) |
| 12 months | 100 | 1 (1%) | 2 (0) | 59 | 1 (2%) | 2 (0) |
| 12 months total | 90 | 3 (3%) | 3 (2) | 53 | 2 (4%) | 2 (1) |
| Daycases |  |  |  |  |  |  |
| baseline | 204 | 62 (30%) | 2 (1) | 98 | 30 (31%) | 2 (3) |
| 6 months | 141 | 42 (30%) | 2 (1) | 66 | 18 (27%) | 1 (1) |
| 12 months | 100 | 17 (17%) | 2 (1) | 59 | 12 (20%) | 2 (2) |
| 12 months total | 90 | 34 (38%) | 2 (2) | 53 | 22 (42%) | 2 (2) |
| **Home Adaptations** |  |  |  |  |  |  |
| baseline | 204 | 40 (20%) |  | 98 | 17 (17%) |  |
| 6 months | 141 | 28 (20%) |  | 66 | 8 (12%) |  |
| 12 months | 100 | 19 (19%) |  | 59 | 9 (15%) |  |

**Supplemental Table 3: complete case descriptive statistics for DeMQoL-U and CarerQoL**

|  | Intervention | | Control |  | Adjusted difference (95% Confidence Interval) |
| --- | --- | --- | --- | --- | --- |
|  | N | mean (SD) | N | mean (SD) |  |
| DeMQoL-proxy utility scores and QALYs | | | | | |
| Baseline utility | 204 | 0.794 (0.095) | 97 | 0.798 (0.092) |  |
| 6 months utility | 150 | 0.758 (0.215) | 69 | 0.747 (0.229) |  |
| 12 months utility | 114 | 0.729 (0.261) | 67 | 0.715 (0.278) |  |
| 12 months QALYs | 104 | 0.753 (0.185) | 60 | 0.747 (0.189) | 0.007 (-0.070 to 0.084) |
| CareQOL | | | | | |
| Baseline CarerQoL | 203 | 77.358 (16.362) | 97 | 77.656 (18.517) |  |
| 6 months CarerQoL | 138 | 80.986 (14.819) | 64 | 79.444 (16.908) | 2.14 (0.15 to 4.12) |
| 12 months CarerQoL | 99 | 81.436 (13.465) | 59 | 80.727 (13.799) | 1.80 (0.51 to 3.10) |
